# Learning Diagnostic Proficiency in Robotic-Assisted Bronchoscopy with Integrated Cone-Beam CT: A 680-Lesion Learning Curve Analysis

**DOI:** 10.64898/2026.07.21.26358556

**Authors:** Nabila Aissami, Carolin Steinack, Reto Engeli, Patrick Baumgartner, Anissa Amstutz, Jacqueline Tanner, Christian Clarenbach, Silvia Ulrich, Malcolm Kohler, Thomas Gaisl

**Affiliations:** Department of Pulmonology, University Hospital Zurich, University of Zurich, Zurich, Switzerland; Department of Internal Medicine, University Hospital Zurich, University of Zurich, Zurich, Switzerland

**Author notes:** **Corresponding author information** Thomas Gaisl, MD PhD MPH University Hospital Zurich Rämistrasse 100 8091 Zurich, Switzerland. Nr.

**Keywords:** Learning Curve, Peripheral pulmonary lesions, robotic-assisted navigation, Cone-beam CT

## Abstract

**Background:** Robotic-assisted bronchoscopy combined with integrated cone-beam computed tomography (RAB+CBCT) enables accurate sampling of peripheral pulmonary lesions (PPLs), but the acquisition of diagnostic proficiency and program-level efficiency remains incompletely characterized.

**Methods:** We conducted a single-center cohort study of consecutive RAB+CBCT (Ion™ endoluminal system, Cios Spin) procedures performed by two experienced interventional pulmonologists. Strict lesion-level diagnostic yield was the primary outcome. Learning curve cumulative sum (LC-CUSUM) analysis determined operator-specific proficiency, followed by conventional CUSUM monitoring of post-proficiency performance. Secondary outcomes included procedure and intubation times, temporal changes in case complexity, and adverse events.

**Results:** Overall, 427 procedures comprising 680 PPLs were analyzed. Median lesion long-axis diameter was 11 mm, 14.4% had a bronchus sign, and 36.5% of procedures involved multiple lesions. Strict lesion-level diagnostic yield was 85.4% (581/680). LC-CUSUM demonstrated proficiency after 48 and 86 PPLs, respectively; thereafter, both operators maintained acceptable performance without crossing the predefined CUSUM decision limit. Median procedure time decreased from 65 minutes during the first 10 procedures to 38 minutes during the last 10. The median intubation time was 70 minutes and declined significantly with increasing experience. Most indicators of lesion complexity remained stable, while short-axis diameter and bronchus-sign prevalence decreased modestly. Adverse-event frequency declined significantly over time.

**Conclusion:** RAB+CBCT achieved high strict diagnostic yield, with heterogeneous operator-specific learning trajectories within a maturing multidisciplinary program. Diagnostic performance, procedural efficiency, and safety improved despite stable or modestly increasing case complexity. These findings support individualized, outcome-based proficiency assessment and longitudinal monitoring, rather than comparative operator ranking or reliance on fixed procedural-volume thresholds.

## Introduction

The increasing use of thoracic CT imaging and implementation of lung cancer screening programs has led to a growing number of peripheral pulmonary lesions (PPLs) requiring diagnostic assessment.(1, 2) The diagnostic performance of conventional bronchoscopy is limited by the requirement for a bronchus sign, navigation inaccuracies, and CT-to-body divergence.(3–5) The combination of robotic-assisted bronchoscopy (RAB) with integrated cone-beam CT (CBCT) enables real-time confirmation of instrument position and has substantially improved navigation success and diagnostic yield while reducing dependence on favorable PPL characteristics.(6–8)

Successful adoption of RAB+CBCT requires mastery of several interconnected workflows, including pre-procedural planning, robotic navigation, intraprocedural imaging, ventilation strategies, radiation management, and tissue acquisition.(9) Beyond operator expertise, implementation depends on coordinated multidisciplinary teamwork, dedicated operating room resources, and substantial institutional investment. Because the procedure is currently performed under general anesthesia, the learning phase may also increase personnel requirements, anesthesia use, operating room occupancy, and associated opportunity costs. Understanding how proficiency, efficiency, and safety evolve during implementation is therefore essential for both clinical outcomes and healthcare resource utilization.(10)

To date, five studies have specifically evaluated clinical adoption for shape-sensing robotic bronchoscopy with or without integrated CBCT.(11–15) Bott et al. reported substantial variability between operators and estimated proficiency after approximately 25 lesions using diagnostic yield as the primary endpoint.(11) Brock et al. analyzed the first European experience and observed improvements in procedure duration after approximately six to eight cases while maintaining high tool-in-lesion rates.(12) More recently, van der Heijden et al. demonstrated stable diagnostic performance from the outset among bronchoscopists already experienced in advanced CBCT-guided navigation bronchoscopy.(13) Zalepurgas et al. demonstrated a learning curve characterized by a progressively improved diagnostic yield. However, existing studies have largely focused on diagnostic yield of single-lesion cases, or technical success (e.g., tool-in-lesion rates) and have been conducted during early adoption phases, with non-integrated CBCT systems, and with relatively limited case numbers.(11–14)

Whether procedural efficiency continues to improve beyond initial adoption remains unclear. This is particularly relevant given the increasing procedural demand and pressure on healthcare resources. Unlike previous studies limited to 50–150 procedures, we evaluated 680 consecutive PPL at a high-volume (>400 procedures/year) European tertiary referral center and applied a prospective, outcome-based proficiency assessment using LC-CUSUM to characterize operator development in procedure duration, workflow efficiency, and procedural performance.

## Methods

### Study design

This single-center cohort study included consecutive patients undergoing RAB using the Ion™ endoluminal system (Intuitive Surgical, Sunnyvale, CA, USA) integrated with mobile cone-beam computed tomography (CBCT; Cios Spin, Siemens Healthineers, Forchheim, Germany) for the diagnosis of PPLs at the University Hospital Zurich between July 2024 and January 2026. PPLs were defined as lesions located beyond the segmental bronchi, without a predefined size threshold, and considered unsuitable for diagnosis using conventional ultrathin bronchoscopy by the investigators. Procedures were performed by two experienced interventional pulmonologists with previous experience in CBCT, electromagnetic navigation, and virtual navigation platforms, and prospectively documented in an institutional registry. The study was approved by the Cantonal Ethics Committee Zurich (BASEC IDs 2024-00684). Data quality was monitored internally and externally.

### Procedural protocol

Procedures were performed under general anesthesia with endotracheal intubation (flexible 8.5 tubes) using a standardized institutional protocol with apneic oxygenation (inspiratory breath holds of up to 20 minutes).(16) In 9% of cases, a lateral decubitus strategy (either primary or secondary) was required, necessitating repositioning of the patient and the entire procedural team, a workflow step that is typically time-consuming.(17) Pre-procedural planning, robotic navigation, and integrated CBCT-guided confirmation of tool-in-lesion were performed as previously described.(9) Radial endobronchial ultrasound (rEBUS) and fluoroscopy were used whenever appropriate. Tissue sampling was performed using combinations of robotic aspiration needles, forceps biopsy, bronchioalveolar lavage (BAL), and transbronchial cryobiopsy (1.1 mm cryoprobe, Erbe Elektromedizin, Thübingen, Germany) at the discretion of the bronchoscopist and according to current guidelines.(18, 19) Endobronchial ultrasound-guided mediastinal staging was performed when clinically indicated.(18)

### Outcomes

The primary outcome was diagnostic yield, defined according to the strict American Thoracic Society (ATS) criteria at the lesion level.(3) Learning curves were evaluated using both learning curve cumulative summation (LC-CUSUM) and cumulative summation (CUSUM) analyses.(13) The secondary outcome was procedural efficiency, assessed by robotic procedure time and total intubation time extracted from the electronic anesthesia record. Lesion-level diagnostic yield was prespecified as the primary endpoint because patients had a mean of 1.6 PPLs, and each lesion required independent planning, navigation, CBCT confirmation, and tissue acquisition. Consequently, each lesion represented a separate opportunity to acquire and assess procedural proficiency, making lesion-level analysis the more appropriate unit for evaluating the learning curve. Additionally, sensitivity analyses were performed at the patient level. Other secondary outcomes included temporal trends in lesion characteristics during program implementation (including axial lesion size, morphology, bronchus sign, pleural distance, and location) and procedural safety at the patient level. Adverse events were classified according to common terminology criteria for adverse events (CTCAE) version 5.0.(20)

### Statistical analysis

Continuous variables are presented as median (interquartile range) or mean±standard deviation, as appropriate, and categorical variables as counts and percentages. Comparisons over time were performed using the Kruskal-Wallis or Mann-Whitney U test for continuous variables and χ² or Fisher’s exact test for categorical variables.

Learning curves for diagnostic yield were assessed using learning-curve cumulative sum (LC-CUSUM) followed by conventional cumulative sum (CUSUM) analysis. LC-CUSUM begins with the assumption that an operator is not yet proficient and sequentially incorporates the outcome of each procedure. Crossing the predefined decision threshold indicates that sufficient statistical evidence has accumulated to classify the operator as proficient. From that point onward, conventional CUSUM analysis was used to determine whether diagnostic performance remained within the predefined acceptable range throughout the remainder of the study. The primary endpoint for both analyses was strict nodule-level diagnostic yield, as defined by the ATS consensus criteria.(3) Decision limits were determined a priori according to the methodology proposed by Biau et al. using predefined acceptable (p₀ = 0.15) and unacceptable (p₁ = 0.25) failure rates, corresponding to acceptable and unacceptable diagnostic yields of 85% and 75% from previous studies, respectively.(6, 21, 22) LC-CUSUM and CUSUM were calculated using the likelihood-ratio method as originally described by Biau et al. and subsequently adapted for RAB learning curve analyses.(21) In addition, conventional CUSUM analyses were performed for robotic procedure duration and total anesthesia/intubation time to evaluate changes in procedural efficiency over the implementation period.

Procedural efficiency was further evaluated using segmented linear regression. Piecewise linear models with a single breakpoint were fitted separately for each operator, and the optimal breakpoint was identified by minimizing the Bayesian Information Criterion (BIC). Regression slopes before and after the breakpoint were used to quantify learning-related improvements and identify the procedural plateau.

To evaluate whether case complexity changed during program implementation, temporal trends in lesion characteristics were analyzed overall and by operator. Continuous variables (short- and long-axis lesion diameter and pleural distance) were assessed using linear regression, whereas binary variables (multiple lesions, bronchus sign, and solid lesion morphology) were analyzed using logistic regression. Models included procedure number as a continuous predictor, and operator-by-time interaction terms were used to assess whether temporal trends differed between operators. Lesion-level analyses accounted for clustering of multiple lesions sampled during the same procedure using cluster-robust standard errors.

Statistical analyses were performed using Stata/MP version 18.0 (StataCorp LLC, College Station, TX, USA) for CUSUM analyses. All tests were two-sided, and p < 0.05 was considered statistically significant.

## Results

Between July 2024 and January 2026, 751 consecutive PPLs were evaluated using RAB with integrated CBCT. After excluding 10 PPLs from patients without informed consent, 18 undergoing concomitant surgical resection, 37 with incomplete primary outcome data, and six that had resolved before the procedure and therefore provided no procedural learning opportunity, the primary learning-curve analysis included 427 procedures targeting 680 PPLs performed by operators A and B. Patient and lesion characteristics are summarized in **Table 1**. No clinically meaningful differences were observed between operators; after false discovery rate correction, only the number of CBCT spins remained significantly different (p < 0.001, q = 0.003), likely reflecting differences in procedural sampling strategy rather than case complexity. Among 680 PPLs, bronchoscopy identified primary lung cancer in 44.4% (302/680), specific benign diagnoses in 30.3% (206/680), and other or metastatic malignancies in 10.7% (73/680), while 13.1% (89/680) were non-diagnostic and 1.5% (10/680) represented technical failures (atelectasis, bleeding, etc.).

**Table 1.** Characteristics of the patients and lesions.

|  | Total | Operator A | Operator B | p-value |
| --- | --- | --- | --- | --- |
| **Patient characteristics** | 427 (100.0%) | 253 (59.3%) | 174 (40.7%) |  |
| Age, years — median [IQR] | 68.3 [61.0-74.6] | 67.8 [61.0-74.4] | 68.7 [61.0-75.0] | 0.498 |
| Male sex — no. (%) | 208 (49.6%) | 123 (49.0%) | 85 (50.6%) | 0.750 |
| Height, cm — median [IQR] | 169 [162-175] | 170 [162-175] | 168 [161-175] | 0.280 |
| Weight, kg — median [IQR] | 71.0 [60.0-83.0] | 73.0 [60.0-84.0] | 70.5 [60.0-81.9] | 0.355 |
| Body mass index, kg/m<sup>2</sup> — median [IQR] | 24.8 [21.9-28.4] | 24.8 [22.0-28.7] | 24.7 [21.6-27.7] | 0.375 |
| Coexisting conditions — no. (%) |  |  |  |  |
| Current or previous cancer | 156 (36.5%) | 92 (36.4%) | 64 (36.8%) | 0.744 |
| Chronic obstructive pulmonary disease | 87 (27.3%) | 43 (23.4%) | 44 (32.6%) | 0.068 |
| GOLD stage 1 | 20 (17.2%) | 8 (14.0%) | 12 (20.3%) |  |
| GOLD stage 2 | 39 (33.6%) | 22 (38.6%) | 17 (28.8%) |  |
| GOLD stage 3 | 23 (19.8%) | 11 (19.3%) | 12 (20.3%) |  |
| GOLD stage 4 | 2 (1.7%) | 1 (1.8%) | 1 (1.7%) |  |
| Unknown | 3 (4.1%) | 1 (1.8%) | 2 (2.4%) |  |
| Oral anticoagulation | 29 (6.8%) | 18 (7.1%) | 11 (6.3%) | 0.605 |
| Antiplatelet therapy | 61 (14.3%) | 42 (16.6%) | 19 (10.9%) | 0.047 |
| Tobacco smoking history |  |  |  | 0.625 |
| Ex-smoker — no. (%) | 169 (40.5%) | 106 (42.4%) | 63 (37.7%) |  |
| Current smoker — no. (%) | 151 (36.2%) | 87 (34.8%) | 64 (38.3%) |  |
| Pack-years among ever smokers — median [IQR] | 40 [20-50] | 40 [20-50] | 40 [25-55] | 0.543 |
| Lung function tests — median [IQR] |  |  |  |  |
| Forced vital capacity, % predicted | 92 [80-105] | 91 [79-100] | 95 [81-109] | 0.030 |
| Forced expiratory volume in 1 second, % predicted | 83 [67-99] | 82 [67-94] | 86 [66-105] | 0.257 |
| Diffusing capacity of the lung for carbon monoxide, % predicted | 73 [56-90] | 71 [58-90] | 76 [55-91] | 0.717 |
| **Lesion characteristics** | 680 (100.0%) | 416 (61.2%) | 264 (38.8%) |  |
| Short axis, mm — median [IQR] | 9 [6-13] | 9 [6-12] | 9 [6-13] | 0.349 |
| Long axis, mm — median [IQR] | 11 [8-17] | 11 [8-16] | 11 [8-17] | 0.549 |
| Bronchus sign — no. (%) | 97 (14.3%) | 51 (12.5%) | 46 (17.4%) | 0.150 |
| Procedures with >1 lesion — n (%) | 248 (36.5%) | 161 (38.7%) | 87 (33.0%) | 0.129 |
| Distance from pleura, mm — median [IQR] | 5 [0-13] | 5 [0-13] | 5 [0-12] | 0.595 |
| Density — no. (%) |  |  |  |  |
| Solid | 388 (57.7%) | 223 (54.5%) | 165 (62.5%) | 0.041 |
| Part-solid | 177 (26.3%) | 118 (28.9%) | 59 (22.3%) | 0.061 |
| Pure ground-glass opacity | 97 (14.4%) | 63 (15.4%) | 34 (12.9%) | 0.363 |
| Concurrent linear EBUS staging — no. (%) | 88 (16.9%) | 49 (15.8%) | 39 (18.6%) | 0.409 |
| Lobes — no. (%) |  |  |  | 0.473 |
| Right upper | 213 (31.3%) | 130 (31.2%) | 83 (31.4%) |  |
| Middle | 43 (6.3%) | 27 (6.5%) | 16 (6.1%) |  |
| Right lower | 141 (20.7%) | 78 (18.8%) | 63 (23.9%) |  |
| Left upper | 169 (24.9%) | 111 (26.7%) | 58 (22.0%) |  |
| Left lower | 114 (16.8%) | 70 (16.8%) | 44 (16.7%) |  |
| CBCT, spins — mean ± SD | 1.32±0.60 | 1.22±0.52 | 1.47±0.68 | <0.001 |
| Fluoroscopy time, s — median [IQR] | 61 [31-102] | 62 [37-103] | 59 [28-102] | 0.202 |
| DAP total, Gy·cm<sup>2</sup> — median [IQR] | 7.0 [4.8–11.5] | 7.1 [4.8–10.9] | 7.0 [4.7–12.1] | 0.277 |
| **Adverse events** | 427 (100.0%) | 253 (59.3%) | 174 (40.7%) |  |
| Any adverse event | 67 (15.7%) | 34 (13.4%) | 33 (19.0%) | 0.111 |
| Bleeding (Nashville grade >1) | 42 (9.8%) | 21 (8.3%) | 21 (12.1%) | 0.235 |
| Respiratory complications (30 days follow-up) | 12 (2.8%) | 6 (2.4%) | 6 (3.4%) | 0.450 |
| Pneumothorax | 13 (3.0%) | 7 (2.8%) | 6 (3.4%) | 0.870 |
| Without chest tube | 6 (1.4%) | 3 (1.2%) | 3 (1.7%) |  |
| Requiring chest tube | 7 (1.6%) | 4 (1.6%) | 3 (1.7%) |  |
| **Procedure times — median [IQR]** | 427 (100.0%) | 253 (59.3%) | 174 (40.7%) |  |
| Preparation (room entry to procedure start), min | 28 [23–35] | 27 [22–34] | 30 [24–39] | 0.005 |
| Procedure (scope-in to scope-out), min | 46 [33–60] | 40 [29–52] | 53 [41–72] | <0.001 |
| Post-procedure (procedure end to extubation), min | 13 [9–19] | 13 [10–18] | 13 [9–19] | 0.93 |
| Other procedural tasks, min* | 2 [0–5] | 2 [0–5] | 2 [0–5] | 0.721 |
| Total intubation time, min | 66 [52–83] | 59 [49–74] | 76 [63–96] | <0.001 |

Values are presented as median [IQR], mean ± SD, or number (n, %) unless otherwise indicated. Percentages may not sum to 100 because of rounding. Lesion diameters were measured in the axial plane. Bronchus sign was defined as CT evidence of a bronchus leading to or traversing the lesion.

**Table 2.** Summary of operator-specific learning curve milestones and procedural performance.

| Outcome | Operator A | Operator B |
| --- | --- | --- |
| Proficiency achieved by LC-CUSUM<sup>1</sup> | 48 PPLs | 86 PPLs |
| Initial procedure time, min<sup>2</sup> | 43 | 65 |
| Final procedure time, min<sup>2</sup> | 38 | 56 |
| Relative reduction | –12% | –18% |
| Initial intubation time, min<sup>2</sup> | 72 | 104 |
| Final intubation time, min<sup>2</sup> | 56 | 70 |
| Relative reduction | –22% | –33% |
| Overall strict lesion-level diagnostic yield | 88.9% (370/416) | 79.9% (211/264) |
| Overall strict patient-level diagnostic yield | 94.1% (238/253) | 87.9% (153/174) |

Abbreviations: LC-CUSUM, learning-curve cumulative sum; min, minutes; PPL, peripheral pulmonary lesion.
<sup>1</sup> Procedural proficiency was determined by learning curve cumulative sum (LC-CUSUM) analysis using strict diagnostic yield at the lesion level as the performance endpoint.
<sup>2</sup> Initial and final procedure and intubation times were estimated from locally weighted scatterplot smoothing (LOWESS) curves using a bandwidth of 0.4 and represent the beginning and end of the learning curve.

### Primary outcomes

Overall, the strict lesion-level diagnostic yield was 85.4% (581/680), comprising 88.9% (370/416) for operator A and 79.9% (211/264) for operator B. LC-CUSUM analysis demonstrated operator-specific acquisition of procedural proficiency, which was achieved after 48 consecutive procedures for operator A and 86 procedures for operator B. Following proficiency, conventional CUSUM analysis confirmed sustained diagnostic performance above the predefined acceptable failure rate for both operators without crossing the decision limit (**Figure 1**). Sensitivity analyses using strict patient-level diagnostic yields produced results comparable to those with strict patient-level diagnostic yields, with unchanged conclusions regarding proficiency acquisition and maintenance. These findings demonstrate high overall diagnostic performance while highlighting substantial inter-operator variability in the number of procedures required to achieve proficiency.

**Figure 1.**
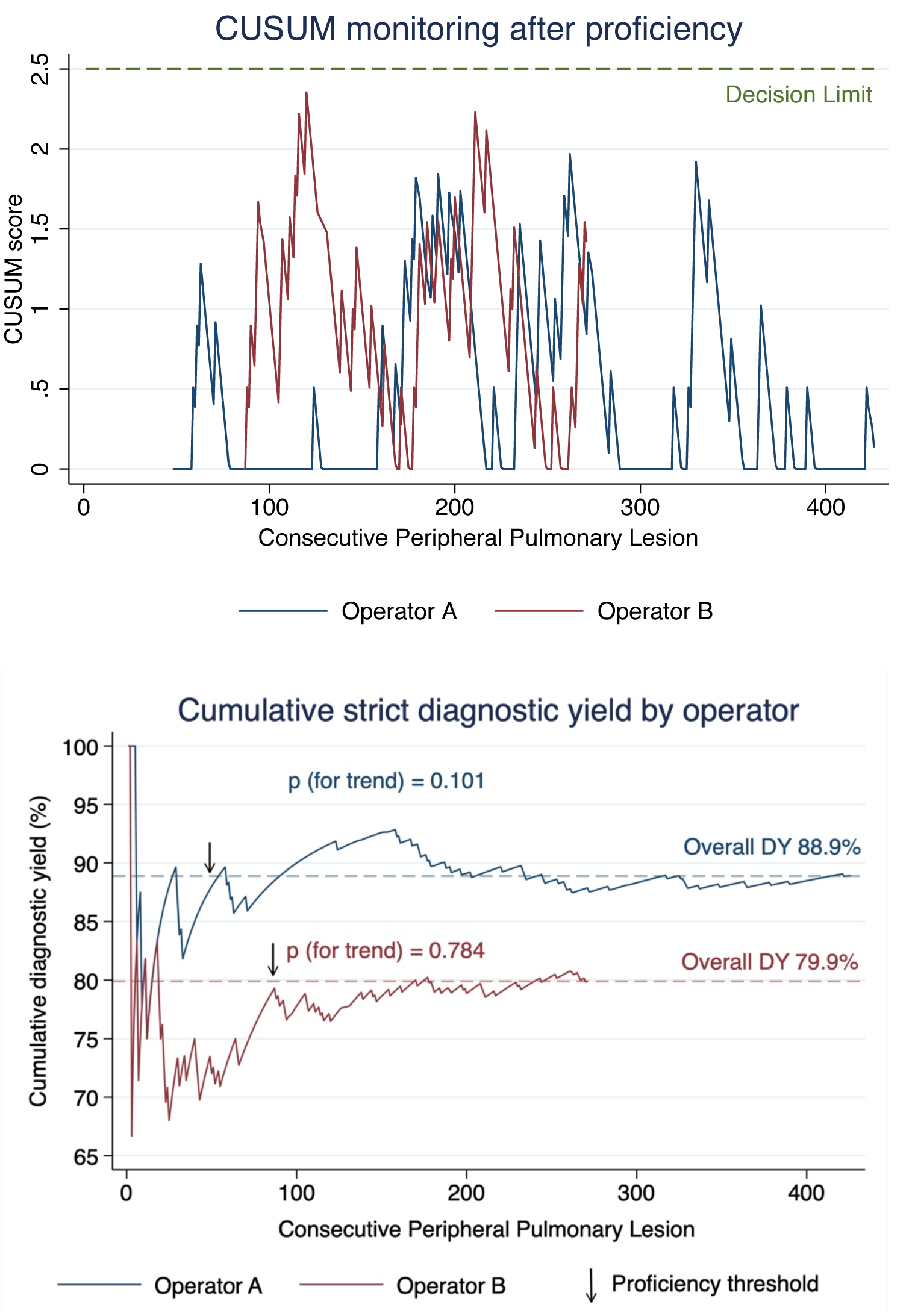
**(A)** LC-CUSUM analysis was first performed under the assumption of non-proficiency to identify when each operator achieved the predefined proficiency threshold. The conventional CUSUM curves shown here begin only after proficiency was established (after 48 consecutive procedures for operator A (blue) and 86 for operator B (red)) and therefore represent post-proficiency quality monitoring using strict lesion-level diagnostic yield. Neither curve crossed the predefined decision limit, indicating sustained acceptable performance throughout the subsequent observation period. **(B)** Cumulative strict lesion-level diagnostic yield for each operator over all consecutive peripheral pulmonary lesion procedures. After initial fluctuations during the learning phase, diagnostic yield stabilized with increasing procedural experience.

#### Secondary outcomes

The median procedure time was 46 [IQR 33–60] minutes across 427 procedures (680 PPLs). Procedure time decreased significantly across consecutive procedural deciles (Kruskal– Wallis, p < 0.001) and from 65 minutes during the first 10 procedures to 38 minutes during the last 10 procedures (**Figure 2**). At the operator level, procedure time was inversely correlated with procedural experience for operator B (Spearman’s ρ = −0.282, p = 0.0003), but not for operator A (ρ = −0.100, p = 0.124). The mean intubation time was 69.9 ± 24.2 minutes, 47% longer than the mean procedure time, and similarly decreased across learning phases (Kruskal–Wallis, p < 0.001) with significant inverse correlations for both operators (operator A: ρ = −0.194, p = 0.012; operator B: ρ = −0.440, p < 0.001), the latter demonstrating a steeper decline (**Figure 2**).

**Figure 2.**
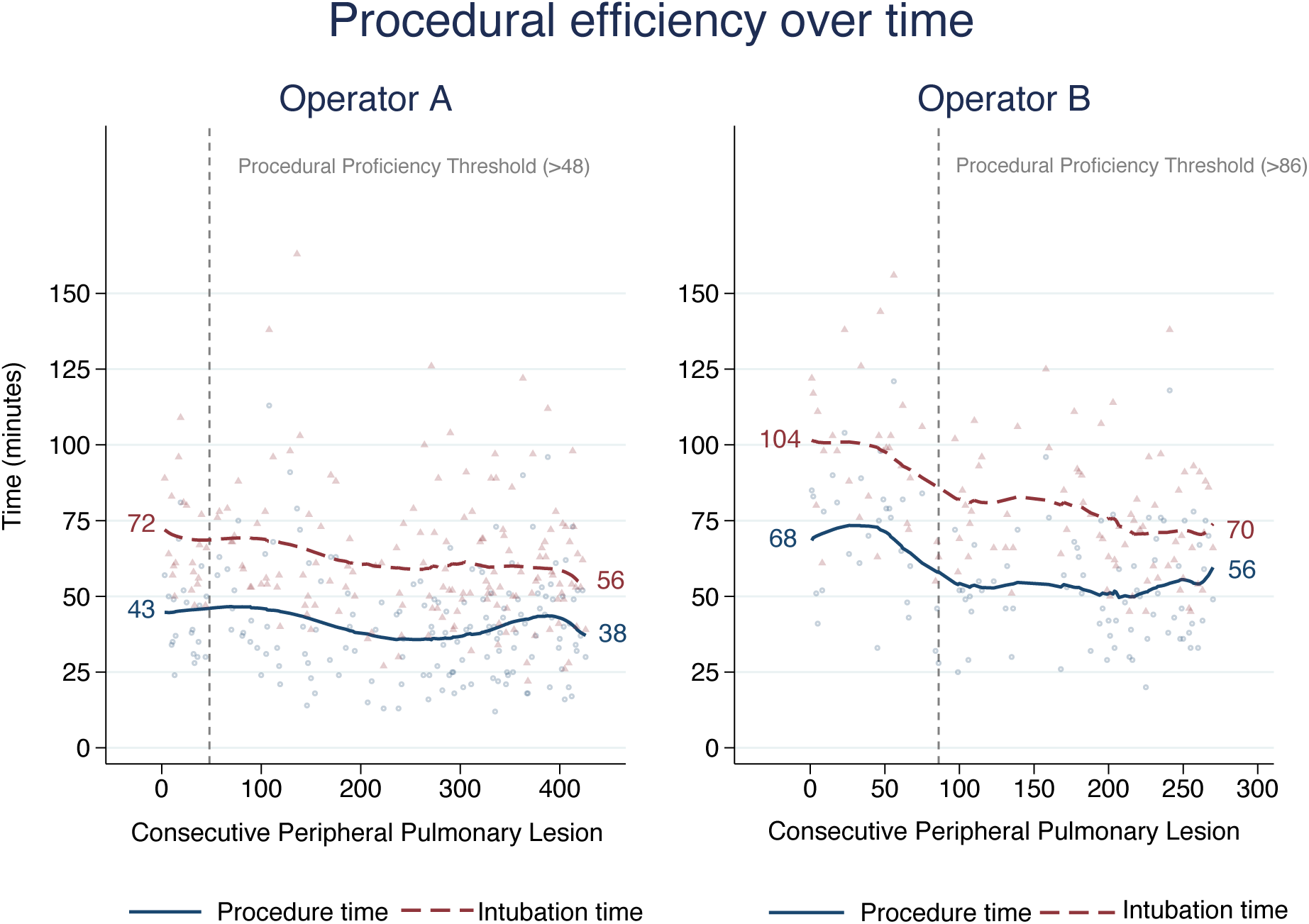
Procedure and intubation times decreased with increasing experience, with the greatest reductions observed before proficiency was achieved. Thereafter, procedure times plateaued, while intubation times continued to decline modestly. Note: Intubation time was measured from tube insertion to removal, and procedure time from bronchoscope insertion to removal, as documented in anesthesia records. Procedure times are reported per patient and include sampling of a mean of 1.6 peripheral lesions, linear endobronchial ultrasound staging in 17%, and lateral decubitus positioning in 9% of procedures.

### Temporal trends

Overall, lesion complexity remained largely stable throughout the implementation period. There were no significant temporal changes in long-axis PPL diameter (overall: β = −0.94 mm across the learning curve, p=0.450), pleural distance (β = −1.66 mm, p=0.225), lesion solidity (overall trend: OR 1.10, p=0.829), the proportion of procedures with multiple target lesions (all p>0.14), or operator-specific temporal trends for these variables (all interaction p>0.45). Short-axis PPL diameter decreased modestly over time (β = −3.13 mm, p=0.002), driven primarily by operator B (β = −0.020 mm per procedure, p=0.003), whereas no significant trend was observed for operator A (β = −0.004 mm per procedure, p=0.153; interaction p=0.082). Likewise, the prevalence of a bronchus sign decreased modestly over time (OR 0.34, 95% CI 0.12–0.96, p=0.042), although no significant operator-by-time interaction was observed (p=0.505). Collectively, these findings suggest that improvements in diagnostic yield and procedural efficiency were unlikely to be explained by major changes in case selection, although the modest reduction in short-axis lesion diameter indicates that later cases were, if anything, slightly more technically challenging.

### Safety profile

Overall, 67 adverse events occurred in 427 procedures, with 62 patients (14.5%) experiencing 1 adverse event and 5 patients (1.2%) experiencing 2 adverse events. Bleeding was the most frequent complication (9.8%, 42/427), followed by respiratory complications, pneumothorax requiring or not requiring chest tube drainage, and airway complications (see Table 1). Cardiac complications, anesthesia-related complications, myocardial infarction, respiratory failure, and hemoptysis were rare (≤1.0% each), while no cases of ARDS, air embolism, hemorrhagic stroke, pulmonary edema, ischemic stroke, or post-procedural fever were observed. According to CTCAE, 46 (68.7%) adverse events were grade 1–2, 19 (23.9%) were grade 3, two (3.0%) were grade 4, and no grade 5 events occurred. The overall incidence of adverse events decreased significantly throughout the implementation period. In logistic regression adjusted for operator, each additional consecutive procedure was associated with a lower likelihood of any adverse event (OR 0.997, 95% CI 0.994–0.999, p=0.005). There was no significant difference in adverse event rates between operators (OR 0.83, 95% CI 0.46–1.51, p=0.542), and the temporal reduction was comparable between operators (interaction p=0.363). These findings suggest that procedural safety improved during the learning phase despite stable case complexity.

## Discussion

In this large single-center cohort of 680 consecutive RAB+CBCT cases, we identified a procedural maturation in diagnostic performance, accompanied by progressive improvements in procedural efficiency and safety. The overall strict diagnostic yield was 85.4%, with both operators crossing the prespecified LC-CUSUM proficiency boundary and maintaining acceptable performance thereafter. Improvements occurred despite largely unchanged lesion complexity throughout the study period, supporting true procedural learning rather than progressive case selection. The observed learning curves likely reflect combined operator and program maturation. Together, these findings support the concept that successful implementation of RAB+CBCT is a longitudinal process that involves not only the acquisition of robotic navigation skills but also the optimization of imaging, sampling, anesthetic, and team workflows.

In our cohort, diagnostic yield improved by approximately 10% during the implementation phase. Such an improvement has been estimated to reduce time to diagnosis by approximately one month, increase survival by 0.36 years, and improve net monetary benefit by approximately US$8,700 per patient.(23) Consequently, shortening the learning phase may have clinically meaningful consequences beyond procedural efficiency.

The overall diagnostic yield compares favorably with previous learning-curve series. Bott et al. reported a yield of 72% across 551 lesions sampled by 9 operators, with substantial interoperator variability and only 6 achieving proficiency during the observed period.(11) By contrast, the present analysis used a strict lesion-level definition of diagnostic yield and predefined an acceptable yield of 85%, thereby applying a more demanding performance standard. The observed 85.4% yield is also consistent with the 85.3% reported in the recent Bonn cohort and with contemporary series combining shape-sensing navigation and intraprocedural three-dimensional imaging.(14) These findings reinforce the importance of distinguishing diagnostic yield from technical endpoints such as navigation success or tool-in-lesion. Although CBCT-confirmed tool-in-lesion is a prerequisite for reliable sampling, it does not guarantee representative tissue acquisition, appropriate specimen handling, or a definitive pathological diagnosis. From a patient and healthcare-system perspective, strict diagnostic yield therefore remains the most clinically meaningful measure of procedural success.

The number of procedures required to establish proficiency was greater than that reported by some previous studies. Bott et al. identified an aggregated change point at approximately 21–25 lesions among operators who ultimately achieved competency, whereas Brock et al. reported immediate technical competence and tool-in-lesion proficiency after approximately eight cases.(11, 12) More recently, Bruinen et al. reported LC-CUSUM proficiency after 42 and 43 procedures, respectively, in two experienced bronchoscopists using CBCT-guided navigation.(13) Direct comparisons are challenging because these studies assessed different outcomes, applied different thresholds for acceptable and unacceptable failure rates, and included substantially different patient and lesion populations. Notably, our cohort comprised relatively small targets, with a median PPL diameter of 10 mm. In particular, technical proficiency, as defined by tool-in-lesion, is expected to be achieved earlier than that defined by strict diagnostic yield.(13) Furthermore, LC-CUSUM assumes initial non-proficiency and requires accumulation of sufficient statistical evidence before crossing its decision boundary; it therefore estimates the point at which proficiency can be declared with predefined certainty rather than the first case at which acceptable clinical performance occurred. The 48- and 86- case thresholds in our study should consequently not be interpreted as minimum credentialling numbers applicable to all bronchoscopists, but as operator-specific estimates under a deliberately stringent 85% acceptable-yield target.

The observed variation in learning trajectories supports outcome-based competency assessment rather than reliance on procedural volume alone. Proficiency reflects the combined development of multiple skills, including registration, navigation, CBCT interpretation, catheter stabilization, sampling strategy, bleeding control, and coordination with anesthesia and technical staff, which may mature at different rates. Brock et al. similarly observed divergent learning patterns across individual procedural components: one operator improved predominantly in procedure time and CBCT utilization, whereas the other improved in registration, rEBUS visualization, and biopsy duration.(12) Bott et al. also found that several operators did not meet competency criteria despite comparable procedural exposure.(11) These observations argue against a single case-number threshold for certification and instead favor structured training with assessment of discrete competencies, followed by longitudinal monitoring of clinically meaningful outcomes. High-volume exposure remains important, but volume should be regarded as an opportunity to acquire competence rather than proof that competence has been achieved.

The procedural-time analyses provide a complementary perspective. Diagnostic proficiency and procedural efficiency are related but distinct domains: an operator may obtain adequate diagnostic results while still requiring excessive time, imaging, or anesthetic support. In our cohort, robotic procedure duration and total intubation time improved over successive cases and demonstrated operator-specific patterns of stabilization. This extends previous reports, which focused mainly on robotic docking-to-undocking time or selected components of the procedure. Bott et al. observed a reduction in median procedure time from 62 minutes during the first 10 cases to 39 minutes after case 40, whereas Brock et al. observed an early reduction after approximately 6 cases.(11, 12) The Bonn cohort similarly showed a decrease from 49 minutes in the early phase to approximately 30 minutes in later phases.(14) Our inclusion of total intubation time is clinically relevant because it captures the broader procedural ecosystem, including anesthetic induction, positioning, airway management, CBCT preparation, team coordination, and emergence. Accordingly, observed efficiency gains should not be attributed exclusively to the bronchoscopist; they reflect maturation of the entire program.

The temporal analysis of lesion characteristics strengthens the interpretation that the observed improvements are consistent with procedural learning. Long-axis diameter, pleural distance, lesion solidity, and the frequency of multiple-target procedures remained stable. Short-axis diameter decreased by approximately 3.1 mm across the study period, and the probability of a positive bronchus sign also declined. These changes would conventionally be expected to reduce rather than increase diagnostic yield. Bott et al. similarly observed that operators increasingly selected smaller lesions with less favorable rEBUS views and absent bronchus signs after their initial learning phase.(11) Such evolution is characteristic of a maturing referral program: once confidence increases, operators expand indications to lesions previously considered less suitable for bronchoscopic sampling. Therefore, the maintenance or improvement of performance despite increasing technical difficulty provides stronger evidence of skills acquisition than improvement observed in progressively easier cases.

Safety also improved during implementation. Bleeding was the most frequent adverse event, while pneumothorax, respiratory complications, airway complications, and severe cardiopulmonary events were uncommon. The odds of any adverse event decreased significantly with increasing procedural experience, without a significant operator-by-time interaction. This temporal reduction is compatible with improved catheter control, biopsy positioning, sampling strategy, hemostasis, and peri-procedural coordination. Nevertheless, the overall adverse-event frequency should be interpreted in light of comprehensive prospective capture and the inclusion of low-grade CTCAE events, which produce higher rates than studies reporting only serious complications. The absence of grade 5 events and the rarity of grade 4 complications remain reassuring.

### Limitations

Several limitations should be considered. The retrospective, single-center, two-operator design limits external validity. Both operators were experienced interventional pulmonologists with prior navigational bronchoscopy experience in a high-volume program supported by dedicated anesthesia, nursing, radiology, and industry expertise; consequently, the learning curves reported here may underestimate those encountered in lower-volume centers or among operators without advanced bronchoscopy experience. Procedural techniques and sampling strategies evolved over time, making operator- and program-level learning inseparable. Because diagnostic-yield classification was based primarily on the ATS strict consensus statement, incorporating clinical and radiological follow-up could allow some initially non-diagnostic benign or nonspecific results to be reclassified as diagnostic for either operator.(3) Furthermore, operator-specific differences should not be interpreted as direct comparative measures of individual clinical competence. Because case allocation was not randomized, residual differences in lesion characteristics, temporal exposure, sampling strategy, procedural assistance, pathology classification, and evolving team workflows may have contributed to the observed trajectories. With only two operators, individual and program-level effects cannot be separated reliably. Finally, CUSUM analyses are inherently sensitive to the predefined acceptable and unacceptable failure rates and decision thresholds.(24)

### Conclusion

In conclusion, RAB with integrated CBCT achieved a high diagnostic yield, with sustained performance after operator-specific learning periods. Improvements in diagnostic performance, procedural efficiency, and safety occurred despite stable or modestly increasing case complexity. These findings support outcome-based monitoring of procedural proficiency and suggest that competency in RAB should be defined by clinical performance rather than procedural volume alone.

## Author contributions

Study concept and design: N.A., R.E., T.G. Acquisition of data: N.A., R.E., P.B., A.A., J.T., T.G. Analysis and interpretation of data: All authors. Drafting of the manuscript: N.A., T.G. Critical revision of the manuscript for important intellectual content: All authors. Statistical analysis: N.A., R.E., T.G. Administrative, technical, or material support: C.S., R.E., C.C., S.U., M.K., T.G. Study supervision: S.U., M.K., T.G.

## Data availability statement

The datasets supporting the findings of this study are not publicly available due to patient privacy and institutional data protection requirements, but are available from the corresponding author upon reasonable request.

## Acknowledgements

Grammarly (Grammarly Inc., San Francisco, CA, USA) was used solely for minor English language and grammar corrections. ChatGPT (OpenAI, GPT-5.5) was used to assist with manuscript organization, language editing, and proofreading. Neither tool was used to generate scientific content, interpret data, perform analyses, or influence the scientific conclusions. All outputs were critically reviewed, verified, and revised by the authors, who take full responsibility for the content of the manuscript.

